# shinyCurves, a shiny web application to analyse multisource qPCR amplification data: a COVID 19 case study

**DOI:** 10.1101/2020.10.14.20212381

**Authors:** S. Olaechea-Lázaro, I. García-Santisteban, JR. Pineda, I. Badiola, S. Alonso, JR. Bilbao, N. Fernandez-Jimenez

## Abstract

**Summary:** Quantitative, reverse transcription polymerase chain reaction (qRT-PCR) has been the gold-standard tool for viral detection during the SARS-CoV-2 pandemic. However, the desperate rush for a quick diagnosis led the use of very different types of machines and proprietary software, leading to an unbearable complexity of data analysis with a limited parameter setup. Here, we present shinyCurves, a shiny web application created to analyse multisource qPCR amplification data from independent multi-plate format. Furthermore, our automated system allows the classification of the results as well as the plot of both amplification and melting curves. Altogether, our web application is an automated qPCR analysis resource available to the research community.

**Availability:** The shinyCurves web application to analyze multisource qPCR amplification data is publicly available under CC license (CC BY-NC-SA 4.0) at https://biosol.shinyapps.io/shinycurves/ and https://github.com/biosol/shinyCurves.

## 1. Introduction

Quantitative, reverse transcription polymerase chain reaction (qRT-PCR) is a widely used technique for the detection and quantification of mRNA. Currently, with the spread of Covid-19, this technique has become the gold-standard tool for SARS-CoV-2 detection (World Health Organization (WHO), 2020). Thus, depending on the amount of viral mRNA detected, a sample is assigned as “Positive”, “Negative” or “Undetermined”. Most healthcare providers rely on automatized detection procedures like the COBAS 6800 system (Hoffmann-La Roche) that makes use of a proprietary software and offers limited parameter setting-up (or control).

In this context, following WHO recommendations, research laboratories world-wide are developing alternative diagnostic protocols that do not depend on commercial kits and their accompanying software. Thus, researchers need to choose from a wide variety of both quantitative gene expression reagents (one-*vs*. two-step qRT-PCR or fluorescent probes *vs*. intercalating dyes) and adapt protocols to the real-time amplification systems available (Guruceaga *et al*., 2020). In addition to the need for independent experimental protocols, there is also a need for an open-source tool that can analyze qRT-PCR data irrespective of the protocol or equipment used. Additionally, in order to discard samples with unspecific amplification products, a software for accurate qRT-PCR analysis should allow the inspection of amplification and melting curves.

Herein we present shinyCurves, a Shiny-based, user-friendly application that is able to process qRT-PCR amplification raw data obtained with either fluorescent probes or intercalating dyes in different qRT-PCR systems. Specifically, when attempting to detect the presence of circulating viral nucleic acids, the application allows the user to decide the settings that will classify samples into three categories (“Positive”, “Negative” or “Undetermined”). Besides, shinyCurves offers the possibility to plot both amplification and melting curves, providing additional quality control on the specificity of the amplification. With the hope that our application can be useful for many laboratories in the current pandemic, we provide shinyCurves as a free resource applicable to the detection of viral infections. A Covid-19 toy dataset is also provided as a practical example.

## 2. Implementation

shinyCurves is designed for users with limited or no programming experience who wish to analyse qRT-PCR data in a simple and efficient manner. The application was completely written using R (R Core Team, 2020) in combination with the ‘shiny’ package (Chang *et al*., 2020), and can be found in the shinyapps.io repository as a web application (https://biosol.shinyapps.io/shinycurves/). The source code can be freely downloaded from the GitHub repository https://github.com/biosol/shinyCurves. Analysis tables are processed by ‘data.table’ (Dowle and Srinivasan, 2018) and ‘dplyr’ (Wickham *et al*., 2020) due to their high efficiency, and plots are generated using ‘ggplot2’ (Wickham, 2016) and ‘plotly’ (Sievert, 2020). Specifically, to plot melting curves the ‘qpcR’ R package (Spiess, 2018) is invoked.

Raw data can be uploaded directly in different file formats: csv, xlsx or xls generated in two widely used qPCR systems, in particular the BioRad and Applied Biosystems platforms (Figure 1). shinyCurves allows for the use of sample duplicates and is also independent of the plate format, i.e. 96- or 364-well plates. The inclusion of experimental controls and serial dilutions of viral DNA is optional, but if included, it must follow a specific format (see Manual).

**Figure 1.**
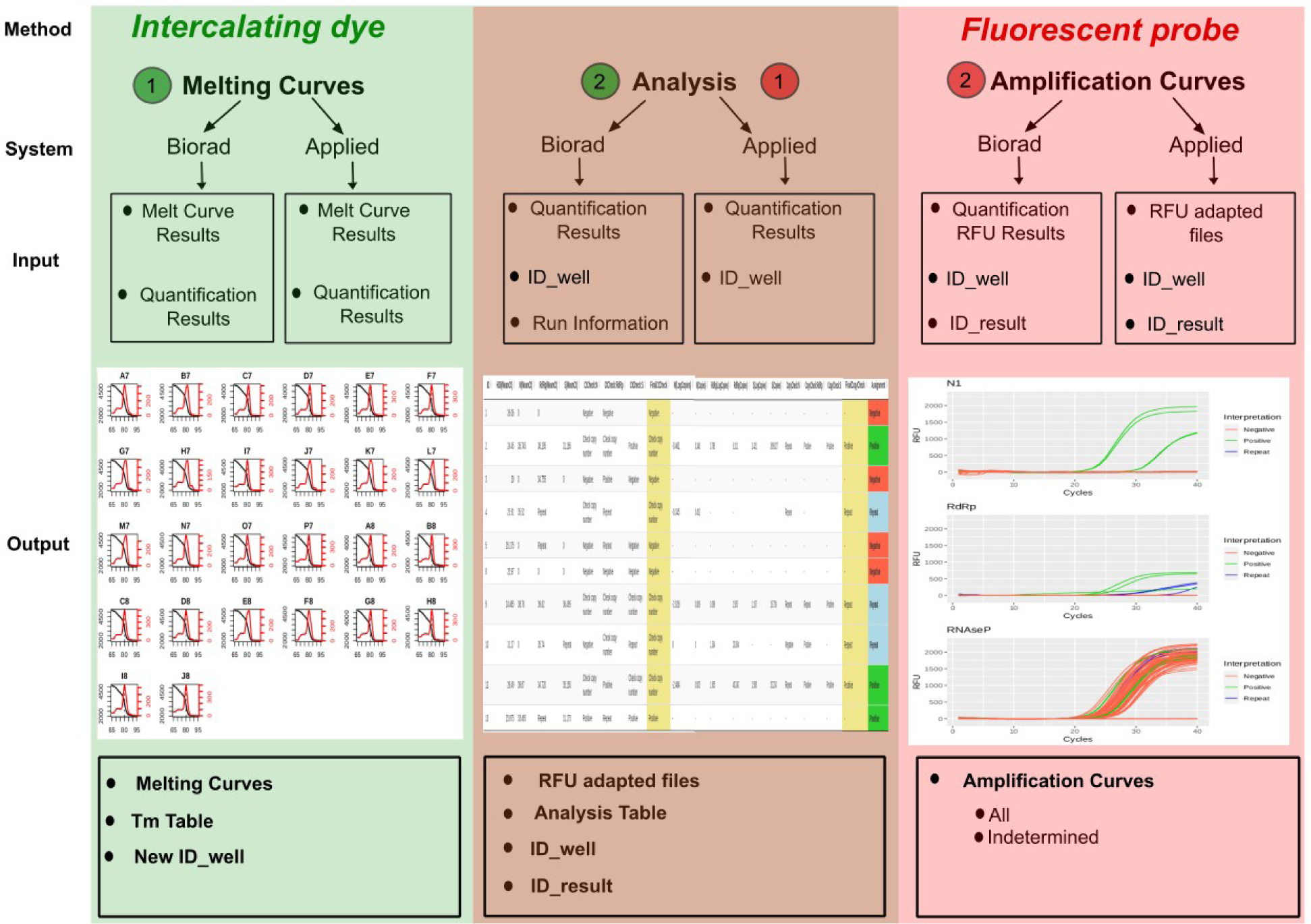
An example overview of two shinyCurves analyses (intercalating dye and fluorescent probe, in green and pink, respectively). Both analyses can be performed using qRT-PCR results from Biorad CFX or Applied Biosystems Quant Studio platforms, among others. The input data required for each analysis are specified in the upper black boxes. In brown, the calling analysis, which follows an initial melting curve analysis in intercalating dye experiments, or is followed by an amplification curve analysis in case fluorescent probes have been used.

## 3. Results

To illustrate the shinyCurves pipeline, data sets generated in the COVID-19 Basque Inter-Institutional Group (coBIG) coming both from fluorescent probes and intercalating dye experiments run on BioRad CFX and Applied Biosystems Quant Studio systems are provided (Guruceaga *et al*., 2020). Genes chosen in these analyses are *N1, RdRp* and *RNAseP* (endogenous control) in the probe assay and *N, S, RdRp* and *H30* (endogenous control) in the dye assay. These genes are included in the Covid-19 diagnostic panel described by the US Centers for Disease Control and Prevention (CDC, https://www.cdc.gov/).

### 3.1 Melting curves (in intercalating dye analysis)

In the dye experiments, shinyCurves allows users to plot melting curves with the aim of excluding from the final calling those samples that do not show reliable and unique melting temperature (Tm) peaks. These plots are generated using the *meltcurve* function from the ‘qpcR’ package in R (Spiess, 2018). As a result, users can download a table including only those plate wells that contain samples with unique Tm peaks that meet the established criteria, together with the Tm value and plot assigned to each well. This table must be included in the calling analysis as a prior filtering step (*ID_well* file).

### 3.2 Calling analysis

After uploading the input data, users are allowed to fine-tune a series of parameters that will be used to call a sample as “Positive”, “Negative” or “Undetermined”. In general, two types of calling criteria are allowed, namely, the Ct value of the analyzed viral gene(s) (compulsory) and the estimated viral RNA copy number (optional). A human endogenous control is included to make sure that the nucleic acid extraction worked. Adjustable parameters include among others: use of a viral DNA standard curve (yes/no), use of duplicates (yes/no) and number of “Positive” viral genes to consider a sample “Positive”. Calling results are presented in a downloadable table. For more details on the calling criteria, see Supplementary Figure 1.

### 3.3 Amplification curves (in fluorescent probe analysis)

After performing the calling analysis in probe experiments, shinyCurves allows to plot General Amplification Curves that include all samples and individual Amplification Curves of “Undetermined” samples. Each “Undetermined” sample is plotted independently, together with those with a final calling. Upon visual inspection of “Undetermined” sample curves, the user can decide whether these fit a sigmoidal distribution, i.e, the result of a specific amplification, or not.

## 4. Discussion

shinyCurves is a user-friendly application that analyses and allows the visualization of qRT-PCR data coming from different amplification methods and platforms. It is easily accessible for any user profile, as no programming skills are required. shinyCurves is set up automatically in shinyapps.io server, making basic Internet connection its only requirement. To our knowledge, this is the first tool designed to automatize the calling of clinical samples containing pathogenous nucleic acids (such as nasopharyngeal samples in COVID-19 testing) through qRT-PCR, and that is completely flexible as regards the user’s requirements and experimental settings. shinyCurves allows a significant improvement in analytical capacity, speed and reproducibility, which are always key factors in pathogen detection analyses, especially in Covid-19 times.

## Supporting information

Manual

## Data Availability

The shinyCurves web application to analyze multisource qPCR amplification data is publicly available under CC license (CC BY-NC-SA 4.0) at https://biosol.shinyapps.io/shinycurves/ and https://github.com/biosol/shinyCurves.

## Acknowledgments

We would like to thank coBIG & “Acción especial COVID-19” participating scientists and voluntaries for the COVID-19 samples and material. This work was supported by CONV20/05 program.

## Supplementary figure legend

**Supplementary Figure 1**. Result assignment criteria in the Calling Analysis. In the upper section, samples are assigned a result based on their viral gene Ct values (compulsory). In the blue section, samples are assigned a result based on their estimated viral DNA copy number (optional). This second analysis is run only for those samples called as “Check copy number” in the Ct Check Analysis.

