## Supplementary material for "shinyCurves, a shiny web application to analyse multisource qPCR amplification data: a COVID 19 case study": Manual

### shinyCurves

#### Index


##### Case-study: Covid-19

a) Fluorescent probes - Applied Biosystems Quant Studio
- b.1) Calling analysis
- b.2) Amplification Curves

b) Intercalating dye - BioRad
- b.1) Melting Curves
- b.2) Calling analysis

##### Manual

0) Before running your analysis

1) Analysis choice

2) File input and parameter choice
- a) Load Data
  - a.1) BioRad CFX
  - a.2) Applied Biosystems Quant Studio
- b) Parameter choice

3) Melting Curves (Intercalating dye)
- a) Load Data
  - a.1) BioRad CFX
  - a.2) Applied Biosystems Quant Studio
- b) Parameter choice
- c) Melting Curves

4) Calling analysis
- a) Fluorescent probes/Intercalating dye Analysis

5) Amplification Curves (Fluorescent probes)
- a) Load Data
- b) Amplification Curves

##### References

### Case-study: **COVID-19**

The toy dataset provided in shinyCurves consists of real RT-qPCR data from clinical nasopharyngeal samples suspicious of containing SARS-CoV-2.
For the PROBE analysis, we have included data from an Applied Biosystems Quant Studio system and for the DYE Analysis data come from BioRad CFX system.

#### a) PROBE - Applied Biosystems Quant Studio

In this dataset, 3 genes (*N1*, *RdRp* and *RNAseP*) will be analyzed in 59 samples. Duplicates are included. *RNAseP* is the endogenous control and *N1* and *RdRp* are the target genes.

##### a.1) Calling analysis

a.1.1) Open the app and click on PROBE: Analysis - Applied Biosystems Quant Studio check box. Upload file “PROBE_Applied Biosystems_384w_Quantification Cq Results.*xlsx*” from the toy dataset folder. This file is located in the **PROBE-Applied/input** folder.

a.1.2) Leave all settings in *Default* mode:

- Endogenous control: *RNAseP*
- Maximum cycle number for endogenous control: 35
- How many serial dilutions are you using for the standard curves?: 4
- Enter standard concentration: 400000
- Sample is considered “Positive” with Ct below: 35
- What is your maximum cycle number: 40
- Number of “Positive” genes to consider a sample "Positive”: 1
- Do you want to use the estimated copy number as a result assignation criterion?: Yes
- Sample is considered "Positive” with estimated copies above: 4

a.1.3) Let’s start by checking that everything is correct in the **Raw Data** tab. The first rows of this tab must look like this:

| Well | Well_Position | Cycle | Fluorescence |
|---|---|---|---|
| 1 | A01 | 1 | -29.2630149929537 |
| 2 | A02 | 1 | 32.1013393571538 |
| 3 | A03 | 1 | 32.7932388987324 |
| 4 | A04 | 1 | 35.0091443249789 |
| 5 | A05 | 1 | -73.0914247778146 |
| 6 | A06 | 1 | 4.3218328184671 |
| 7 | A07 | 1 | 6.4893398141362 |
| 8 | A08 | 1 | 0.355744983633031 |
| 9 | A09 | 1 | 1.26639860236173 |
| 10 | A10 | 1 | 36.8367491872364 |
| 11 | A11 | 1 | -0.267735424668444 |
| 12 | A12 | 1 | 3.5456471434245 |
| 13 | A13 | 1 | 2.04843493278713 |
| 14 | A14 | 1 | 4.46438667400344 |
| 15 | A15 | 1 | 0.358013929156641 |

a.1.4) The **Conversion** tab represents the only difference between the BioRad CFX and the Applied Biosystems Quant Studio Analysis. Usually, RFU amplification data from Applied Biosystems Quant Studio and BioRad CFX are stored in a different formats, so they need to be adapted in order to posteriorly be able to run the **Amplification Curves Analysis**.

In the Applied Biosystems xlsx file, amplification data are stored in the **Raw Data** tab. shinyCurves will read through the tab and generate 3 independent CSV files with the amplification data specific for the wells of each gene. Three independent subtabs for the RFU data for *N1*, *RdRp* and *RNAseP* will appear: download each of those files, since they will be needed when running the **Amplification Curves Analysis**.
These files are called *N1.csv*, *RdRp.csv* and *RNAseP.csv* and can be found in the **PROBE-Applied Biosystems/output** folder. Filenames must not be modified.

a.1.5) Let's move to the **Run Information** tab, which should look like this:

| Block Type | 384-Well Block |
|---|---|
| Calibration Background is expired | No |
| Calibration Background performed on | 07-16-2019 |
| Calibration Pure Dye ABY is expired | No |
| Calibration Pure Dye ABY performed on | 07-16-2019 |
| Calibration Pure Dye CY5 is expired | No |
| Calibration Pure Dye CY5 performed on | 07-16-2019 |
| Calibration Pure Dye FAM is expired | No |
| Calibration Pure Dye FAM performed on | 07-16-2019 |
| Calibration Pure Dye JUN is expired | No |
| Calibration Pure Dye JUN performed on | 07-16-2019 |
| Calibration Pure Dye MUSTANG PURPLE is expired | No |
| Calibration Pure Dye MUSTANG PURPLE performed on | 07-16-2019 |
| Calibration Pure Dye NED is expired | No |
| Calibration Pure Dye NED performed on | 07-16-2019 |
| Calibration Pure Dye ROX is expired | No |

a.1.6) The **Applied Results** tab shows the equivalent to the **Raw Data** column in BioRad CFX Analysis, i.e. the adapted format of Applied Biosystems Quant Studio results. It should look like this:

| Well | ID | Target | Fluor | Cq |
|---|---|---|---|---|
| A01 | 1 | N1 | FAM | NA |
| A02 | 9 | N1 | FAM | NA |
| A03 | 17 | N1 | FAM | NA |
| A04 | 25 | N1 | FAM | NA |
| A05 | 33 | N1 | FAM | NA |
| A06 | 41 | N1 | FAM | NA |
| A07 | 49 | N1 | FAM | NA |
| A08 | 57 | N1 | FAM | NA |
| A09 | 1 | RdRp | FAM | NA |
| A10 | 9 | RdRp | FAM | 39.33 |
| A11 | 17 | RdRp | FAM | NA |
| A12 | 25 | RdRp | FAM | NA |
| A13 | 33 | RdRp | FAM | NA |
| A14 | 41 | RdRp | FAM | NA |
| A15 | 49 | RdRp | FAM | NA |
| A16 | 57 | RdRp | FAM | NA |
| A17 | 1 | RNAseP | FAM | 23.2661910663516 |

Make sure that names for NTC, negative control and dilutions are defined correctly. For example,

| | | | | |
|---|---|---|---|---|
| I08 | N1_10-5 | N1 | FAM | NA |
| O24 | RNAseP_negC | RNAseP | FAM | 23.3931005405676 |
| G08 | NTC_N1 | N1 | FAM | NA |

a.1.7) The **Ct Plate** shows the Ct values for each sample in the plate order. Blank spaces in the plate represent NA values in the Ct column of the raw data, and therefore, empty wells in the experimental plate.

Ct Plate

| | 1 | 2 | 3 | 4 | 5 | 6 | 7 | 8 | 9 | 10 | 11 | 12 | 13 | 14 | 15 | 16 | 17 | 18 | 19 | 20 | 21 | 22 | 23 | 24 |
|---|---|---|---|---|---|---|---|---|---|---|---|---|---|---|---|---|---|---|---|---|---|---|---|---|
| A | | | | | | | | | | 39.330 | | | | | | | 23.266 | 23.679 | 23.344 | 24.013 | 25.349 | | | |
| B | | | | | | | | | | 39.130 | | | | | | | 23.165 | 23.984 | 23.295 | 24.165 | 25.450 | | | |
| C | 34.900 | | | | | | | | 38.000 | | | | | | | | 22.557 | 26.260 | 23.082 | 25.125 | 23.862 | | | |
| D | 35.100 | | | | | | | | 38.500 | | | | 36.089 | | | | 22.418 | 26.462 | 23.008 | 25.280 | 24.015 | | | |
| E | | | | | | 36.089 | | | | | | | | | | | 25.257 | 26.094 | 24.576 | 27.331 | 25.823 | | | |
| F | | | | | | 36.200 | | | | | | | | | | | 25.095 | 26.640 | 24.686 | 27.081 | 25.899 | | | |
| G | | | | | | | | | | | | | | | | | 23.739 | 27.010 | 24.883 | 25.027 | | | | |
| H | | | | | | | | | | | | | | | | | 23.743 | 27.052 | 25.202 | 24.661 | | | | |
| I | | | | | | | | | | | | | | | | | 24.971 | 25.994 | 23.726 | 25.001 | | | | |
| J | | | | | | | | | | | | | | | | | 24.962 | 26.109 | 23.878 | 25.014 | | | | 24.907 |
| K | | | | | | | | 36.320 | | | | | | | | 35.834 | 26.057 | 27.192 | 21.484 | 25.069 | | | | 4.245 |
| L | | | | | | | | 36.992 | | | | | | | | 33.723 | 26.261 | 27.256 | 21.853 | 25.148 | | | | 3.180 |
| M | | | | | | | | 32.742 | | | | | | | | 30.597 | 23.840 | 23.965 | 26.053 | 24.703 | | | | 3.716 |
| N | | | | | | | | 32.377 | | | | | | | | 31.329 | 23.837 | 24.079 | 25.951 | 25.020 | | | | 4.230 |
| O | | 23.953 | | | | | | 29.690 | | 22.596 | | | | | | 27.212 | 23.321 | 25.464 | 24.029 | 24.326 | | | | 23.393 |
| P | | 23.680 | | | | | | 29.561 | | 23.960 | | | | | | 26.774 | 23.382 | 25.282 | 24.167 | 24.560 | | | | 23.868 |

Showing 1 to 16 of 16 entries    Previous 1 Next

a.1.8) Similarly to the Ct Plate, the **Sample Plate** tab represents the samples in the plate set up and should look like this:

Sample Plate

| | 1 | 2 | 3 | 4 | 5 | 6 | 7 | 8 | 9 | 10 | 11 | 12 | 13 | 14 | 15 | 16 | 17 | 18 | 19 | 20 | 21 | 22 | 23 | 24 |
|---|---|---|---|---|---|---|---|---|---|---|---|---|---|---|---|---|---|---|---|---|---|---|---|---|
| A | 1 | 9 | 17 | 25 | 33 | 41 | 49 | 57 | 1 | 9 | 17 | 25 | 33 | 41 | 49 | 57 | 1 | 9 | 17 | 25 | 33 | 41 | 49 | 57 |
| B | 1 | 9 | 17 | 25 | 33 | 41 | 49 | 57 | 1 | 9 | 17 | 25 | 33 | 41 | 49 | 57 | 1 | 9 | 17 | 25 | 33 | 41 | 49 | 57 |
| C | 2 | 10 | 18 | 26 | 34 | 42 | 50 | 58 | 2 | 10 | 18 | 26 | 34 | 42 | 50 | 58 | 2 | 10 | 18 | 26 | 34 | 42 | 50 | 58 |
| D | 2 | 10 | 18 | 26 | 34 | 42 | 50 | 58 | 2 | 10 | 18 | 26 | 34 | 42 | 50 | 58 | 2 | 10 | 18 | 26 | 34 | 42 | 50 | 58 |
| E | 3 | 11 | 19 | 27 | 35 | 43 | 51 | 59 | 3 | 11 | 19 | 27 | 35 | 43 | 51 | 59 | 3 | 11 | 19 | 27 | 35 | 43 | 51 | 59 |
| F | 3 | 11 | 19 | 27 | 35 | 43 | 51 | 59 | 3 | 11 | 19 | 27 | 35 | 43 | 51 | 59 | 3 | 11 | 19 | 27 | 35 | 43 | 51 | 59 |
| G | 4 | 12 | 20 | 28 | 36 | 44 | 52 | NTC_N1 | 4 | 12 | 20 | 28 | 36 | 44 | 52 | NTC_RdRp | 4 | 12 | 20 | 28 | 36 | 44 | 52 | NTC_RNAseP |
| H | 4 | 12 | 20 | 28 | 36 | 44 | 52 | NTC_N1 | 4 | 12 | 20 | 28 | 36 | 44 | 52 | NTC_RdRp | 4 | 12 | 20 | 28 | 36 | 44 | 52 | NTC_RNAseP |
| I | 5 | 13 | 21 | 29 | 37 | 45 | 53 | N1_10-5 | 5 | 13 | 21 | 29 | 37 | 45 | 53 | RdRp_10-5 | 5 | 13 | 21 | 29 | 37 | 45 | 53 | NTC_RNAseP |
| J | 5 | 13 | 21 | 29 | 37 | 45 | 53 | N1_10-5 | 5 | 13 | 21 | 29 | 37 | 45 | 53 | RdRp_10-5 | 5 | 13 | 21 | 29 | 37 | 45 | 53 | NTC_RNAseP |
| K | 6 | 14 | 22 | 30 | 38 | 46 | 54 | N1_10-4 | 6 | 14 | 22 | 30 | 38 | 46 | 54 | RdRp_10-4 | 6 | 14 | 22 | 30 | 38 | 46 | 54 | |
| L | 6 | 14 | 22 | 30 | 38 | 46 | 54 | N1_10-4 | 6 | 14 | 22 | 30 | 38 | 46 | 54 | RdRp_10-4 | 6 | 14 | 22 | 30 | 38 | 46 | 54 | |
| M | 7 | 15 | 23 | 31 | 39 | 47 | 55 | N1_10-3 | 7 | 15 | 23 | 31 | 39 | 47 | 55 | RdRp_10-3 | 7 | 15 | 23 | 31 | 39 | 47 | 55 | |
| N | 7 | 15 | 23 | 31 | 39 | 47 | 55 | N1_10-3 | 7 | 15 | 23 | 31 | 39 | 47 | 55 | RdRp_10-3 | 7 | 15 | 23 | 31 | 39 | 47 | 55 | |
| O | 8 | 16 | 24 | 32 | 40 | 48 | 56 | N1_10-2 | 8 | 16 | 24 | 32 | 40 | 48 | 56 | RdRp_10-2 | 8 | 16 | 24 | 32 | 40 | 48 | 56 | RNAseP_negC |
| P | 8 | 16 | 24 | 32 | 40 | 48 | 56 | N1_10-2 | 8 | 16 | 24 | 32 | 40 | 48 | 56 | RdRp_10-2 | 8 | 16 | 24 | 32 | 40 | 48 | 56 | RNAseP_negC |

Showing 1 to 16 of 16 entries    Previous 1 Next

a.1.9) The Standard Curves tab shows **a Summary Table** and the **Standard Curve plots**. Target genes have amplified in dilutions 10-2, 10-3 and 10-4. Since no amplification was detected in dilution 10-5, standard curves will have only 3 points (instead of 4).

| | Dilution | Copies | logCopies | N1(Dup1) | N1(Dup2) | N1(Avg) | N1(LogCopies) | N1(Copies) | RdRp(Dup1) | RdRp(Dup2) | RdRp(Avg) | RdRp(LogCopies) | RdRp(Copies) |
|---|---|---|---|---|---|---|---|---|---|---|---|---|---|
| NTC | NA | NA | NA | NA | NA | NA | NA | NA | NA | NA | NA | NA | NA |
| C(-) | NA | NA | NA | NA | NA | NA | NA | NA | NA | NA | NA | NA | NA |
| 10-2 | 100.00 | 4000.00 | 3.60 | 29.69 | 29.56 | 29.63 | 3.54 | 3454.69 | 27.21 | 26.77 | 26.99 | 3.61 | 4060.16 |
| 10-3 | 1000.00 | 400.00 | 2.60 | 32.74 | 32.38 | 32.56 | 2.71 | 514.40 | 30.60 | 31.33 | 30.96 | 2.59 | 388.00 |
| 10-4 | 10000.00 | 40.00 | 1.60 | 36.32 | 36.99 | 36.66 | 1.56 | 36.01 | 35.83 | 33.72 | 34.78 | 1.61 | 40.63 |
| 10-5 | 100000.00 | 4.00 | 0.60 | NA | NA | NA | NA | NA | NA | NA | NA | NA | NA |

| RNAseP(Dup1) | RNAseP(Dup2) | RNAseP(Avg) | RNAseP(LogCopies) | RNAseP(Copies) |
|---|---|---|---|---|
| NA | NA | NA | NA | NA |
| 23.39 | 23.87 | 23.63 | 23.63 | 427070919440743457095680.00 |
| NA | NA | NA | NA | NA |
| NA | NA | NA | NA | NA |
| NA | NA | NA | NA | NA |
| NA | NA | NA | NA | NA |

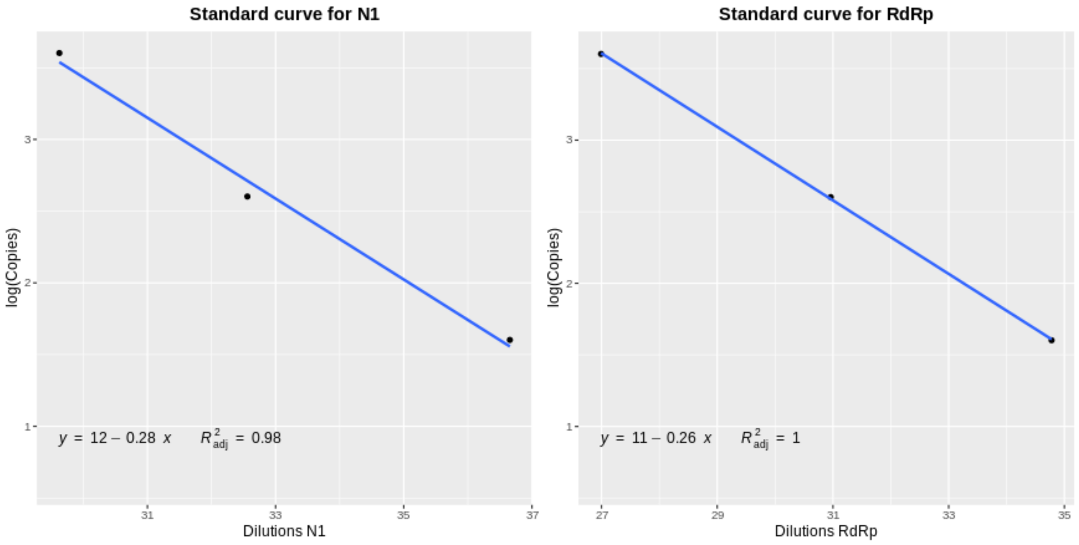


a.1.10) The **Analysis tab** shows the main results for all the samples. This table can be found in CSV format as *Analysis.csv* in the toy dataset (**PROBE-Applied /output** folder).

Remember that in this analysis the **estimated copy number will be used** as result assignation criterion (non-mandatory).

In the following section, the result assignation criteria for the most relevant samples will be described:

<u>Sample 1</u>

This sample shows no amplification neither for *N1* (N1(MeanCt) = 0) nor for *RdRp* (RdRp(MeanCt) = 0), so both genes are assigned as “Negative” in columns *CtCheck:N1* and *CtCheck:RdRp*. As a consequence, the sample can be directly assigned as “Negative” in the *FinalCtCheck* and in the *Assignment* columns.

In this case, there is no need to perform the “*Copy Check*” analysis.

This example is representative for samples 3-8, 10-15, 17-33, 35-42 and 44-59.

<u>Sample 2</u>

This sample shows a mean Ct of of 38.25 for *RdRp and* is “Undetermined” for *N1* due to differences between duplicates. *N1* is assigned as “Undetermined” in its *FinalCtCheck* column and *RdRp* is assigned as “Check copy number” as its Ct value is higher than MaxCt but lower than the maximum cycle number. As a consequence, the sample is assigned as “Check copy number” in the *FinalCtCheck* column and is redirected to the “*Copy Check*” analysis.

Copy number is calculated only for *RdRp* (3.880). As this value is lower than the minimum copy number to assign a sample as “Positive” and *N1* has been assigned as “Undetermined”, this sample is finally assigned as “Undetermined” in the *Assignment* column.

Sample 9

This sample shows a mean Ct of 0 for *N1* and a mean Ct of 39.23 for *RdRp*. *N1* is assigned as "Negative" in its *CtCheck* column (as a Ct value of 0 implies no amplification) and *RdRp* is assigned as "Check copy number" (it is higher than MaxCt).

In this case, the *FinalCtCheck* column assigns the sample as "Check copy number" because, although *N1* is "Negative", *RdRp* could be "Positive" in the "*Copy Check"* analysis.

Copy number for *N1* is 0 (as there is no amplification) and it is assigned as "Negative" in its *CopyCheck* column. On the other hand, *RdRp* shows a value of 2.92 copies, which is lower than #MinCopy (4). For this reason, *RdRp* is assigned as "Undetermined" in its *CopyCheck* column: the copy number is so low, that it cannot be assigned as "Positive" with certainty.

Sample 16

This sample shows a mean Ct of 23.816 for *N1* and of 23.278 for *RdRp*. Both Ct values are lower than MaxCt (35), so that both samples are assigned as "Positive" in their respective *CtCheck* columns and also in the *FinalCtCheck* column. There is no need to perform the "*Copy Check"* analysis, so the sample is directly assigned as "Positive".

Sample 34

In this case, *N1* shows a Ct value of 0 (no amplification) and *RdRp* is assigned as "Undetermined" in its *MeanCt* colum. By looking at the independent Ct values of the duplicates for sample 34 in *RdRp* (NaN and 36.089) we can see that one of them has not amplified and thus, the average cannot be calculated.

As a result, the sample is assigned as "Undetermined" in the *FinalCtCheck* and in the *Assignment* columns: although *N1* is "Negative", we have not been able to perform the "*Ct Check"* analysis for *RdRp* and, thus, the sample cannot be directly assigned as "Negative".

Sample 43

This sample shows a mean Ct value of 36.145 for *N1* and no amplification for *RdRp* (mean Ct = 0). As a consequence, *N1* is assigned as "Check copy number" and *RdRp* as "Negative" in their respective *CtCheck* columns. In this case, the sample is assigned as "Check copy number" in the *FinalCtCheck* column.

In the "*Copy Check"* analysis, *RdRp* shows no copies (it has not amplified) and is again assigned as "Negative". *N1* shows an estimated copy number of 50.179, which is higher than 4 (#MinCopy), and thus, it is assigned as "Positive" as we only need one gene to be "Positive" to consider the sample "Positive".

CAREFUL: if users decide that they **don't want to use the estimated copy number as assignation criteria**, they are allowed to modify this option in the main menu and the Analysis table will be updated instantaneously. However, this could critically modify the results for some samples: users should be aware of this difference!
Specifically for the toy dataset, results would be modified in the following way:

Sample 2
*N1* is assigned as "Positive" as its Ct value = 35 and *RdRp(MeanCt)* = 38.25, so that *RdRp* is assigned as "Negative". The sample is assigned as "Positive" as we only need one gene to be "Positive" to consider the sample "Positive".

Sample 9
With *N1(MeanCt)* = 0 (no amplification) and *RdRp(MeanCt)* = 39.23 (higher than *MaxCt*), both genes are assigned as "Negative" and the sample is finally assigned as "Negative".

Sample 43
With *N1(MeanCt)* = 36.145 (higher than MaxCt) and *RdRp(MeanCt)* = 0 (no amplification), both genes are assigned as "Negative" and the sample is finally assigned as "Negative".

a.1.11) The **ID_Well** tab displays the table (with columns *Well*, *Target*, *ID*) that will later be needed in the **Amplification Curves Analysis.** This file is called *ID_well.csv* and it can be found in the toy dataset in the **PROBE-Applied /output** folder.

a.1.12) The **ID_Result** tab displays a table with *ID* and *Interpretation* columns, which are taken from the *Assignment* column in the central Analysis tab. This file will be needed for the **Amplification Curves Analysis**: it is called *ID_result.csv* and it can be found in the toy dataset in **PROBE-Applied/output** folder. Additionally, it summarizes the results of the main analysis.

#### a.2) Amplification Curves

a.2.1) Click the *Amplification Curves* check box in the menu. Upload the RFU files "*N1.csv*", "*RdRp.csv*" and "*RNAseP.csv*" in the "*Upload CSVs here*" input option. These files must contain the raw RFU data directly exported from RT-qPCR system. They can be found in the toy dataset in folder **PROBE-Applied/output**: select all of them at one time and upload them together.
Remember that filenames must not be modified.

a.2.2) Upload the *ID_well.csv* and *ID_result.csv* files in their respective input options. These files can also be found in the **PROBE-Applied/output** folder in the toy dataset.

a.2.3) Leave the two other settings in Default mode:
- Enter cycle number: 40
- Enter endogenous control: *RNAseP*

a.2.4) Check that **ID_Well** and **ID_Result** tabs are correctly organized! The **ID_Well** tab must display the 3 columns present in the *ID_well.csv* file and the **ID_Result** tab must display the columns *ID* and *Interpretation*, just as in the *ID_result.csv* file.

The headers look like this:

ID_Well

| Well | Target | ID |
|---|---|---|
| A01 | N1 | 1 |
| A02 | N1 | 9 |
| A03 | N1 | 17 |
| A04 | N1 | 25 |
| A05 | N1 | 33 |
| A06 | N1 | 41 |
| A07 | N1 | 49 |
| A08 | N1 | 57 |
| A09 | RdRp | 1 |
| A10 | RdRp | 9 |
| A11 | RdRp | 17 |
| A12 | RdRp | 25 |
| A13 | RdRp | 33 |
| A14 | RdRp | 41 |
| A15 | RdRp | 49 |
| A16 | RdRp | 57 |
| A17 | RNAseP | 1 |
| A18 | RNAseP | 9 |

ID_Result

| ID | Interpretation |
|---|---|
| 1 | Negative |
| 2 | Positive |
| 3 | Negative |
| 4 | Negative |
| 5 | Negative |
| 6 | Negative |
| 7 | Negative |
| 8 | Negative |
| 9 | Repeat |
| 10 | Negative |
| 11 | Negative |
| 12 | Negative |
| 13 | Negative |
| 14 | Negative |
| 15 | Negative |
| 16 | Positive |
| 17 | Negative |
| 18 | Negative |

a.2.5) The **General Plots** tab will display the amplification curves for *N1*, *RdRp* and *RNAseP* for all the samples in different subplots. Samples assigned as “Positive” are plotted in green, samples assigned as “Negative” are plotted in red and samples assigned as “Undetermined” are plotted in blue.

“Positive” samples show usually a sigmoidal amplification curve, while “Negative” samples show usually a plateau curve close to 0.

These plots can be found in *GeneralAmplificationCurves.png* in the **PROBE-Applied/output** folder.

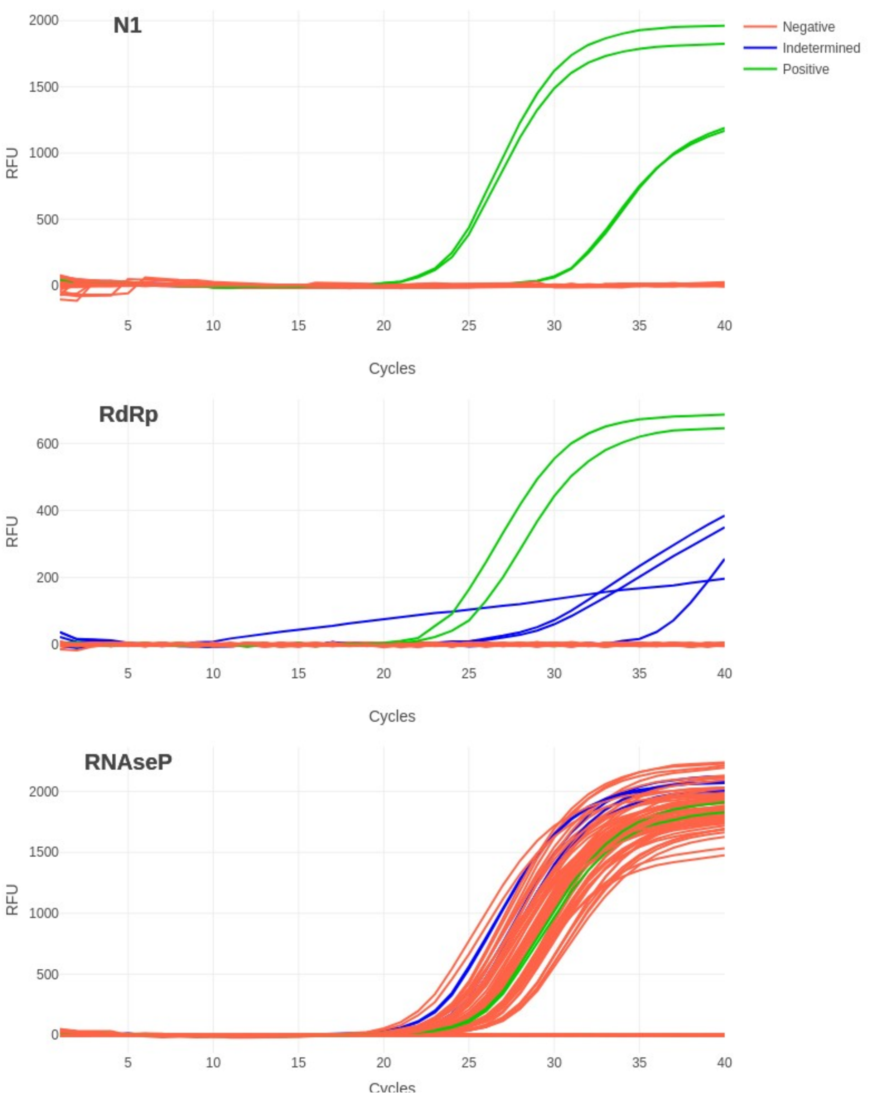


a.2.6) The **Undetermined Samples** tab shows individual amplification curves only for those samples assigned as "Undetermined", plotted against the rest of the samples. Three subtabs are displayed, one for each gene. In this case, samples 9 and 34 are assigned as "Undetermined".

These plots can be found in *N1_IndetAmpCurves.png*, *RdRp_IndetAmpCurves.png* and *RNAseP_IndetAmpCurves.png* files.

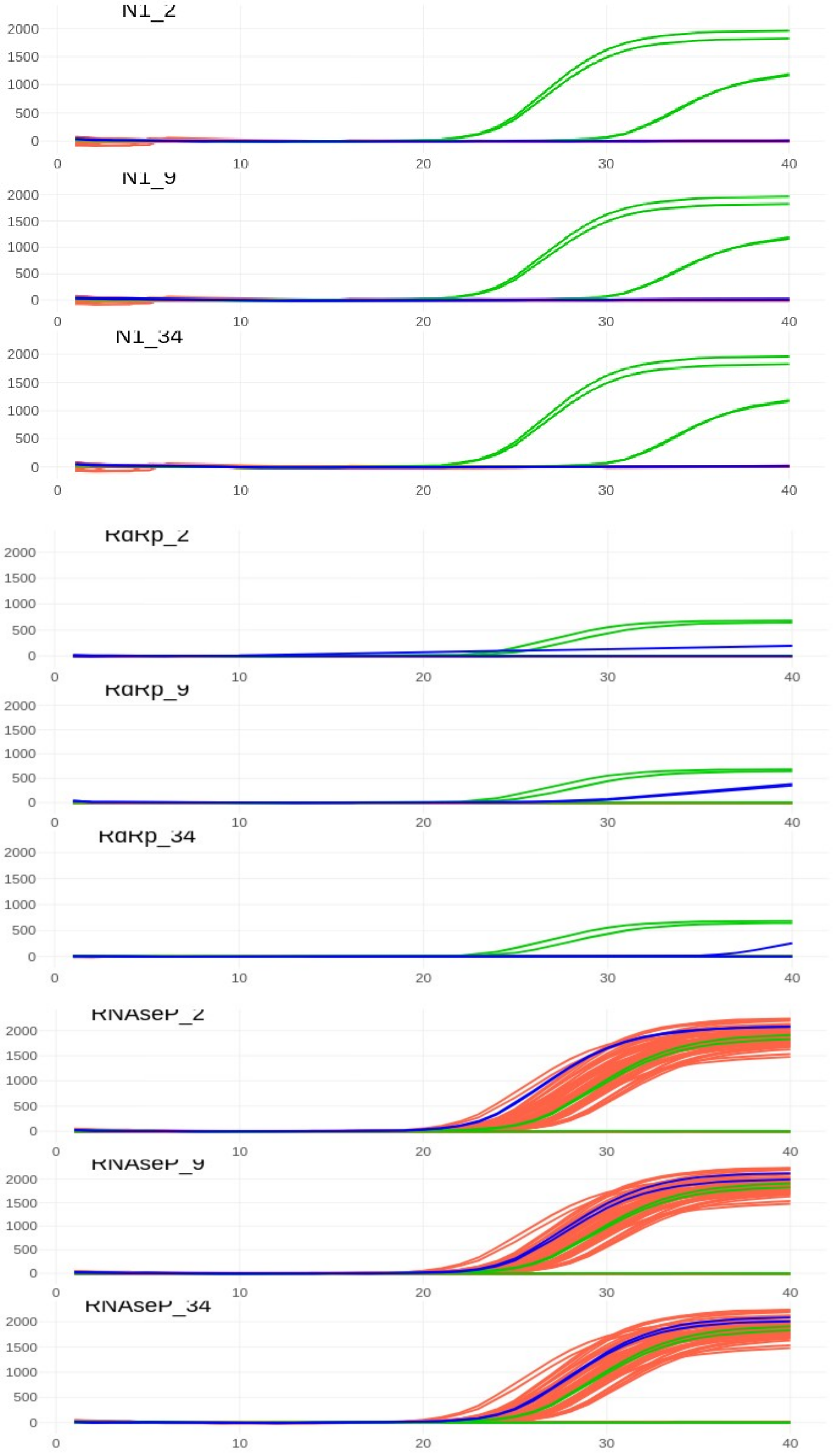

N1_2
N1_9
N1_34
RdRp_2
RdRp_9
RdRp_34
RNAseP_2
RNAseP_9
RNAseP_34
2000
1500
1000
500
0
0
10
20
30
40

#### b) DYE - BioRad CFX

In this dataset, 4 genes (*N*, *S, RdRp* and *H30*) will be analyzed in 7 samples. Duplicates are included. *H30* is the endogenous control and *N, RdRp and S* are the target genes. Different to the PROBE example, this experiment was run in a 96-well plate.

##### b.1) Melting Curves

b.1.1) Click the Melting Curves check box in the main menu and upload files “DYE_BioRad_96w_Melt Curve RFU Results_*H30.csv*”, “DYE_BioRad_96w_Melt Curve RFU Results_*S.csv*”, “DYE_BioRad_96w_Melt Curve RFU Results_*N.csv*” and “DYE_BioRad_96w_Melt Curve RFU Results_*RdRp.csv*” in the “*Upload Melt Curve RFU Results (BioRad/Applied) here*”. Upload all files together at the same time. These files can be found in the **DYE-BioRad/input** folder.

b.1.2) Upload “*DYE_BioRad_96w_Quantification Summary.csv*” file in the “*Upload Quantification Results (BioRad/Applied) here*”. This file can be found in the **DYE-BioRad/input** folder in the toy dataset.

b.1.3) Leave all settings in Default mode:
- Enter cutoff area value: 10
- Enter lower Tm limit: 0.5
- Enter upper Tm limit: 0.5
- Are your data already in first derivative transformed format?: No

b.1.4) The **ID_Well** tab displays a table with columns “*Well*”, “*Target*” and “*ID*”. Check that all the samples are included in the analysis. The header of this tab looks like this:

| Well | Target | ID |
|---|---|---|
| A05 | RdRp | 1 |
| A08 | H30 | 1 |
| B01 | S | 1 |
| B02 | S | 1 |
| B05 | RdRp | 1 |
| B08 | H30 | 1 |
| C01 | N | 1 |
| D01 | N | 1 |
| A01 | N | 2 |
| A03 | S | 2 |
| B03 | S | 2 |
| C05 | RdRp | 2 |
| C08 | H30 | 2 |
| D05 | RdRp | 2 |
| D08 | H30 | 2 |
| G01 | N | 2 |
| C03 | S | 3 |
| D03 | S | 3 |

b.1.5) The **Melting Curves Plots** tab displays 4 subtabs (one for each gene). Each of these should print independent melting curves for each well analyzed.

As we have specified in the options that our data are in raw format, raw fluorescence data are plotted in black and the curve for the first derivative transformed data is plotted in red. The melting peaks identified are marked with a dotted vertical line.

These plots can be found in files *MeltingCurves_H30.pdf*, *MeltingCurves_N.pdf*, *MeltingCurves_RdRp.pdf* and *MeltingCurves_S.pdf* in folder **DYE-BioRad/output** in the toy dataset.

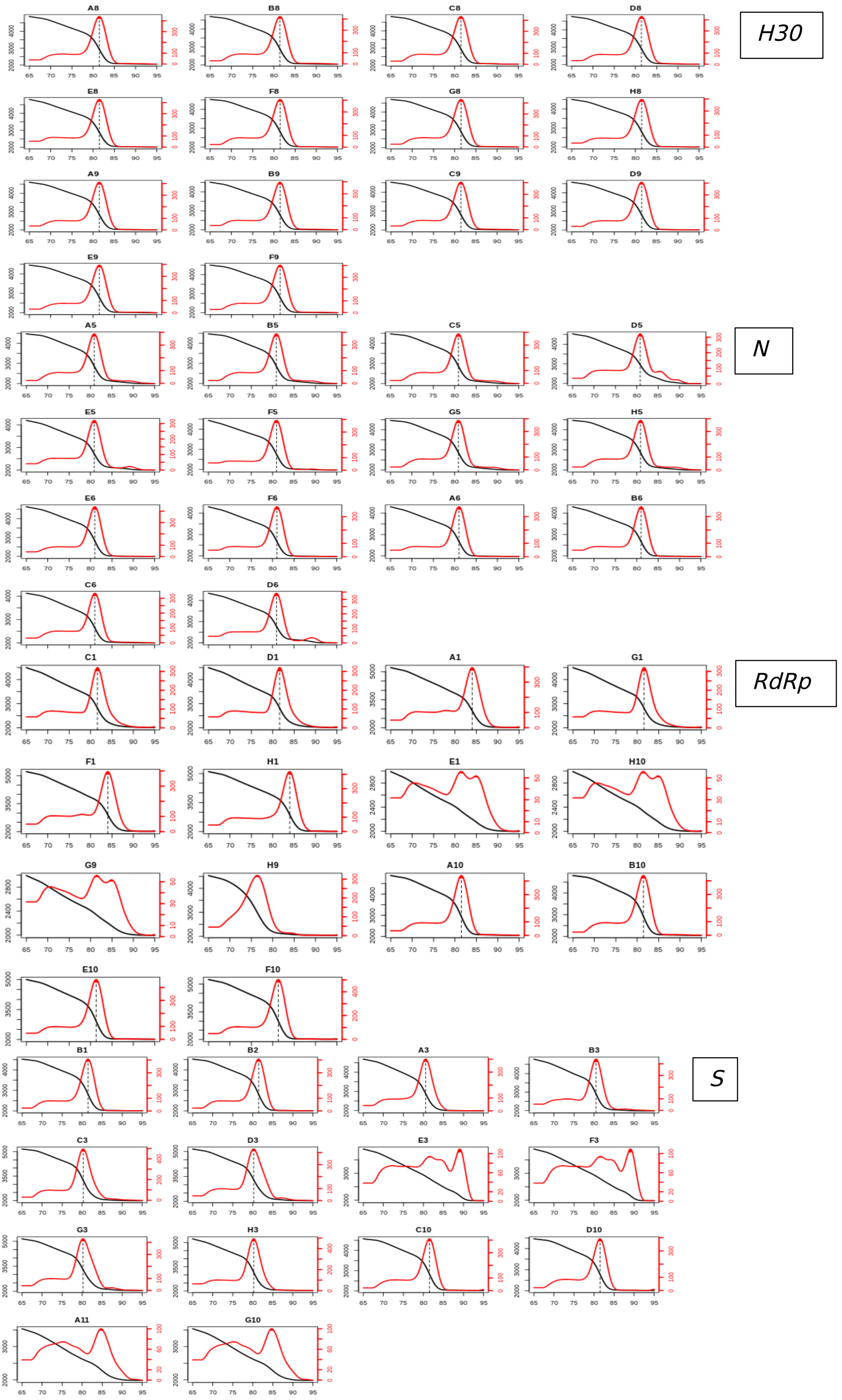

b.1.6) The **Tm Table** displays a table with *ID*, *Well*, *Target*,*Tm* and *Area* only for those Melting Curves that show a unique peak area as specified in the main menu. Wells showing multiple peaks in the Melting Curves plots are considered unreliable (as they might have amplified further than our target genes) and, for this reason, are excluded from this table and will not be included in the central Analysis. This file can be found in the toy dataset in the **DYE-BioRad/output** folder.
*Note: users may check that estimated Tm values do not deviate much from one another or from the expected Tm and discard manually those samples/wells that do.*

The header of this table must look like this:

| ID | Target | Well | Tm | Area |
|---|---|---|---|---|
| 1 | H30 | A8 | 81.45 | 12.57 |
| 1 | H30 | B8 | 81.40 | 11.99 |
| 1 | N | C1 | 81.60 | 10.89 |
| 1 | N | D1 | 81.60 | 10.89 |
| 1 | RdRp | A5 | 80.86 | 12.40 |
| 1 | RdRp | B5 | 80.86 | 12.40 |
| 1 | S | B1 | 81.45 | 11.45 |
| 1 | S | B2 | 81.45 | 11.45 |
| 2 | H30 | C8 | 81.45 | 12.08 |
| 2 | H30 | D8 | 81.40 | 12.02 |
| 2 | N | A1 | 84.01 | 10.94 |
| 2 | N | G1 | 81.60 | 10.89 |
| 2 | RdRp | C5 | 80.86 | 12.40 |
| 2 | RdRp | D5 | 80.81 | 10.07 |
| 2 | S | A3 | 80.57 | 11.71 |
| 2 | S | B3 | 80.47 | 14.12 |
| 3 | H30 | E8 | 81.45 | 12.06 |

b.1.7) The **New ID_Well** tab should display the exactsame table as the Tm Table tab, but only including *ID*, *Target* and *Well* columns. The purpose of this tab is to generate the input file for the main Analysis, showing only wells with reliable melting curves. This file can be found in the toy dataset in the **DYE-BioRad/output** folder.

| ID | Target | Well |
|---|---|---|
| 1 | H30 | A8 |
| 1 | H30 | B8 |
| 1 | N | C1 |
| 1 | N | D1 |
| 1 | RdRp | A5 |
| 1 | RdRp | B5 |
| 1 | S | B1 |
| 1 | S | B2 |
| 2 | H30 | C8 |
| 2 | H30 | D8 |
| 2 | N | A1 |
| 2 | N | G1 |
| 2 | RdRp | C5 |
| 2 | RdRp | D5 |
| 2 | S | A3 |
| 2 | S | B3 |
| 3 | H30 | E8 |
| 3 | H30 | F8 |

#### b.2) Calling analysis

b.2.1) Click the **DYE: Analysis-BioRad** check box in the main menu and upload the file "*DYE_BioRad_96w_Quantification Summary.csv*" in the "*Upload BioRad CFX results file (csv/xlsx/xls) here*". This file can be found in the **DYE-BioRad/input** folder.

b.2.2) Upload *ID_well.csv* file in the "*Upload ID_well here*" input option. This *ID_well.csv* file must be the output obtained from the **New ID_Well** tab in the **Melting Curves Analysis**. This file can be found in the **DYE-BioRad/output** folder.

b.2.3) Leave all settings in the Default mode:
- Endogenous control: *H30*
- Maximum cycle number for endogenous control: 35
- How many serial dilutions are you using for the standard curves?: 4
- Enter standard concentration: 400000
- Sample is considered "Positive" with Ct below: 35
- What is your maximum cycle number: 40
- Number of "Positive" genes to consider a sample "Positive": 2
- Do you want to use the estimated copy number as a result assignation criterion?: Yes
- Sample is considered "Positive" with estimated copies above: 4

b.2.4) Let's start by checking that our *ID_well.csv* file is loaded correctly. Remember that it only contains those wells that have shown a reliable and unique melting curve. The header of this file looks like this:

| ID | Target | Well |
|---|---|---|
| 1 | H30 | A8 |
| 1 | H30 | B8 |
| 1 | N | C1 |
| 1 | N | D1 |
| 1 | RdRp | A5 |
| 1 | RdRp | B5 |
| 1 | S | B1 |
| 1 | S | B2 |
| 2 | H30 | C8 |
| 2 | H30 | D8 |
| 2 | N | A1 |
| 2 | N | G1 |
| 2 | RdRp | C5 |
| 2 | RdRp | D5 |
| 2 | S | A3 |
| 2 | S | B3 |
| 3 | H30 | E8 |
| 3 | H30 | F8 |

b.2.5) The **Run Information** tab displays the message “*No run information to show*”.

Run Information

| data |
|---|
| No run information to show |

b.2.6) The **Raw Data** tab displays those rows of the “*DYE_BioRad_96w_Quantification Summary.csv*” file that correspond to the wells specified in the *ID_well.csv* file and should look like this:

| Well | Fluor | Target | ID | Cq |
|---|---|---|---|---|
| A08 | SYBR | H30 | 1 | 26.00 |
| B08 | SYBR | H30 | 1 | 26.12 |
| C01 | SYBR | N | 1 | 36.42 |
| D01 | SYBR | N | 1 | 35.07 |
| A05 | SYBR | RdRp | 1 | 37.61 |
| B05 | SYBR | RdRp | 1 | 38.76 |
| B01 | SYBR | S | 1 | 31.09 |
| B02 | SYBR | S | 1 | 31.24 |
| C08 | SYBR | H30 | 2 | 24.50 |
| D08 | SYBR | H30 | 2 | 24.40 |
| A01 | SYBR | N | 2 | 38.78 |
| G01 | SYBR | N | 2 | 35.52 |
| C05 | SYBR | RdRp | 2 | 34.28 |
| D05 | SYBR | RdRp | 2 | 35.23 |
| A03 | SYBR | S | 2 | 36.10 |
| B03 | SYBR | S | 2 | 36.89 |

b.2.7) The **Ct Plate** tab must display the following order:

Show 20 entries | Search:

| | 1 | 2 | 3 | 4 | 5 | 6 | 7 | 8 | 9 | 10 | 11 | 12 |
|---|---|---|---|---|---|---|---|---|---|---|---|---|
| A | 38.780 | | 36.100 | 40.100 | 37.610 | | 39.480 | 26.000 | 25.120 | | | |
| B | 31.090 | 31.240 | 36.890 | 39.830 | 38.760 | | | 26.120 | 25.230 | | | |
| C | 36.420 | 34.760 | 39.690 | 33.040 | 34.280 | 35.600 | 35.750 | 24.500 | 25.130 | 31.190 | 32.850 | |
| D | 35.070 | 31.260 | 37.680 | 32.500 | 35.230 | 34.990 | 31.580 | 24.400 | 25.370 | 32.350 | 34.330 | |
| G | 35.520 | 28.870 | 31.090 | 27.670 | | | 26.600 | 25.730 | | | | |
| E | | 32.300 | | 30.390 | | 34.660 | 31.020 | 25.760 | 25.420 | 36.850 | 32.050 | |
| F | 34.180 | 25.850 | | 29.870 | | 34.780 | 30.390 | 26.240 | 25.550 | 34.670 | | |
| H | 32.750 | 29.040 | 31.260 | 27.280 | | | 26.630 | 26.090 | | | | |

Showing 1 to 8 of 8 entries | Previous 1 Next

b.2.8) Similarly, the **Sample Plate** must display the following order:

Show 20 entries | Search:

| | 1 | 2 | 3 | 4 | 5 | 6 | 7 | 8 | 9 | 10 | 11 | 12 |
|---|---|---|---|---|---|---|---|---|---|---|---|---|
| A | 2 | | 2 | S_10-5 | 1 | 6 | RdRp_10-5 | 1 | 5 | 6 | | NTC_H30 |
| B | 1 | 1 | 2 | S_10-5 | 1 | 6 | RdRp_10-5 | 1 | 5 | 6 | | NTC_H30 |
| C | 1 | N_10-4 | 3 | S_10-4 | 2 | 7 | RdRp_10-4 | 2 | 6 | 6 | H30_negC | NTC_N |
| D | 1 | N_10-4 | 3 | S_10-4 | 2 | 7 | RdRp_10-4 | 2 | 6 | 6 | H30_negC | NTC_N |
| G | 2 | N_10-2 | 5 | S_10-2 | | | RdRp_10-2 | 4 | | | NTC_S | NTC_S |
| E | | N_10-3 | | S_10-3 | 3 | 5 | RdRp_10-3 | 3 | 7 | 7 | N_10-5 | NTC_RdRp |
| F | 3 | N_10-3 | | S_10-3 | 3 | 5 | RdRp_10-3 | 3 | 7 | 7 | N_10-5 | NTC_RdRp |
| H | 3 | N_10-2 | 5 | S_10-2 | | | RdRp_10-2 | 4 | | | | |

Showing 1 to 8 of 8 entries | Previous 1 Next

b.2.9) In the **Standard Curve** tab, the Summary Table and the Standard Curve plots should look like this:

| | Dilution | Copies | logCopies | RdRp(Dup1) | RdRp(Dup2) | RdRp(Avg) | RdRp(LogCopies) | RdRp(Copies) | H30(Dup1) |
|---|---|---|---|---|---|---|---|---|---|
| NTC | NA | NA | NA | NA | NA | NA | NA | NA | NA |
| C(-) | NA | NA | NA | NA | NA | NA | NA | NA | 32.85 |
| 10-2 | 100.00 | 4000.00 | 3.60 | 26.60 | 26.63 | 26.62 | 26.62 | 412097519097332064296370176.00 | NA |
| 10-3 | 1000.00 | 400.00 | 2.60 | 31.02 | 30.39 | 30.70 | 30.70 | 5069907082747023631245287358464.00 | NA |
| 10-4 | 10000.00 | 40.00 | 1.60 | 35.75 | 33.58 | 34.66 | 34.66 | 46238102139925934764661647322644480.00 | NA |
| 10-5 | 100000.00 | 4.00 | 0.60 | 39.48 | NA | 39.48 | 39.48 | 3019951720401994409707192832329938632704.00 | NA |

| H30(Dup2) | H30(Avg) | H30(LogCopies) | H30(Copies) | S(Dup1) | S(Dup2) | S(Avg) | S(LogCopies) | S(Copies) | N(Dup1) | N(Dup2) | N(Avg) | N(LogCopies) | N(Copies) |
|---|---|---|---|---|---|---|---|---|---|---|---|---|---|
| NA | NA | NA | NA | NA | NA | NA | NA | NA | NA | NA | NA | NA | NA |
| 34.33 | 33.59 | 1.85 | 71.00 | NA | NA | NA | NA | NA | NA | NA | NA | NA | NA |
| NA | NA | NA | NA | 27.50 | 27.53 | 27.52 | 3.36 | 2275.39 | 25.50 | 25.53 | 25.52 | 4.07 | 11641.73 |
| NA | NA | NA | NA | 32.12 | 31.49 | 31.80 | 2.35 | 224.36 | 30.12 | 29.49 | 29.80 | 3.05 | 1132.69 |
| NA | NA | NA | NA | 36.75 | 34.68 | 35.72 | 1.43 | 27.16 | 34.75 | 32.68 | 33.72 | 2.13 | 135.47 |
| NA | NA | NA | NA | 40.38 | 40.28 | 40.33 | 0.35 | 2.25 | 40.38 | NA | 40.38 | 0.56 | 3.63 |

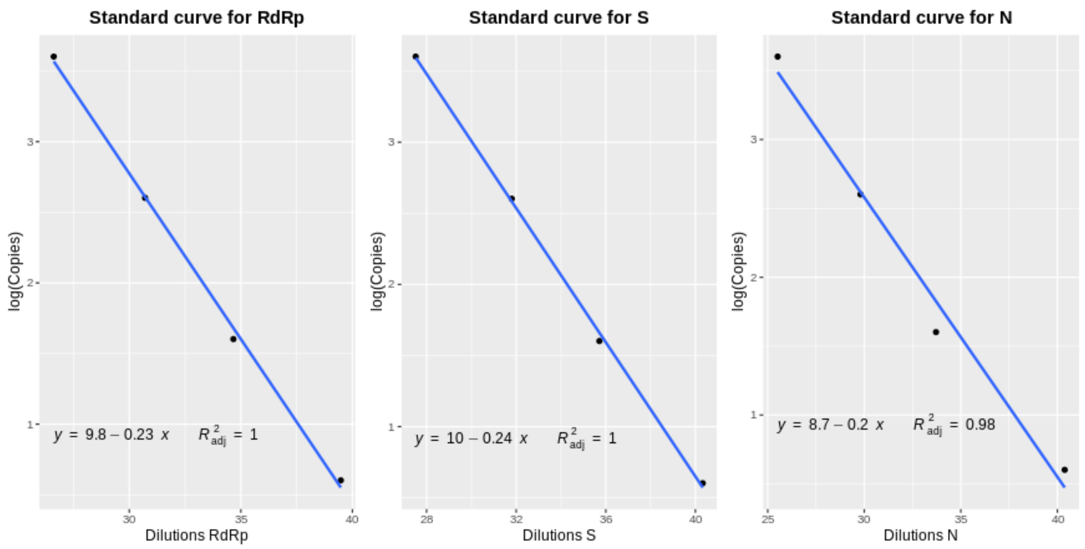


b.2.10) The **Analysis tab** shows the main analysis for all samples. This table can be found in CSV format with name *Analysis.csv* in the toy dataset (in folder **DYE-BioRad/output**).
Recall that, for this analysis, the estimated copy number will be used as a result assignation criteria.

**IMPORTANT**: sample 4 is excluded from the final Analysis. If you take a look at the ID_Well, you will see how this sample has "lost" most of their respective wells in the **Melting Curves Analysis**. Commonly melting curves show multiple peaks due to the absence of specific amplification. However, when the melting curves for all target genes in the same sample look "wrong", i.e. there is no specific amplification for any of the genes (except for the endogenous control), this means that this sample can be assigned as "Negative". If the melting curve for the endogenous control looks unreliable too, this means that the sample has bad quality.
For our analysis, **we need 2 "Positive" genes to assign a sample as "Positive"**. So, as we don't have 2 target genes in sample 4, it is directly excluded from the analysis.

In the following section, the result assignation criteria for the most relevant samples will be described:

Sample 1
For both *N* (N(MeanCt) = 35.745) and *RdRp* (RdRp(MeanCt) = 38.185) this sample shows mean Ct values higher than MaxCt (35) and lower than the RT-qPCR cycle number. As a consequence, both genes are assigned as "Check copy number" in their respective *CtCheck* columns. On the contrary, *S* shows a Ct value of 31.165, which is lower than MaxCt and is, consequently, assigned as "Positive". As 2 of the 3 target genes are assigned as "Check copy number", the whole sample is redirected to the "*Copy Check*" analysis.
Copy number is calculated for the 3 genes: *N*(Copies) = 25.943, *RdRp*(Copies) = 7.156 and *S*(Copies) = 541.155. The three genes show copy numbers higher than #MinCopy (4), so that all of them are assigned as "Positive" in their respective *CopyCheck* columns.
Finally, the sample is assigned as "Positive" in the *FinalCopyCheck* and in the *Assignment* column.

Sample 2

This sample shows a Ct value lower than MaxCt for *RdRp* (RdRp(MeanCt) = 34.755), which is why it is assigned as “Positive” in its *CtCheck* column. For *S*, it shows a Ct value higher than MaxCt but lower than the maximum cycle number so that, consequently, it is assigned as “Check copy number”. *N* is assigned as “Undetermined” due to inconsistencies between sample duplicates. Consequently, the whole sample is assigned as “Check copy number”, as *S* could be “Positive”.

Copy number is calculated for *RdRp* (Copies = 45.614) and *S* (Copies = 29.930) and both show values higher than #MinCopy. As a consequence, the sample is finally assigned as “Positive”.

Sample 3

This sample has not amplified for *RdRp* (MeanCt = 0) and so this gene is assigned as “Negative” in its respective *CtCheck* column. *S* shows unreliable duplicates, so it is assigned as “Undetermined” and *N* shows a Ct value lower than MaxCt (*N*(MeanCt) = 33.465), so it is assigned as “Positive”. This sample is directly assigned as “Undetermined” as we don’t have enough reliable results to make an assignment.

Sample 5

For this sample, *RdRp* (MeanCt = 34.720) and *S* (MeanCt = 31.175) show Ct values lower than MaxCt, so both genes are assigned directly as “Positive”. As a consequence, the whole sample is assigned as “Positive”.

Note that sample 5 has “lost” its wells in the **Melting Curves Analysis** so it is not considered for this analysis.

Sample 6

Neither *RdRp* nor *N* have amplified in this sample (MeanCt = 0) so both genes are assigned as “Negative”. Even if *S* (MeanCt = 31.770) is assigned as “Positive”, the sample is finally assigned as “Negative” because 2 of our 3 target genes are assigned as “Negative”.

Sample 7

Both *RdRp* and *N* are assigned as “Check copy number” as they both show Ct values higher than MaxCt but lower than the RT-qPCR cycle number (*RdRp*(MeanCt = 35.295) and *N*(MeanCt = 39.760)). Copy number is calculated for both genes: while *RdRp* shows a copy number of 34.077 and is assigned as “Positive”, *N* shows a copy number of 3.976 and is assigned as “Undetermined” (the copy number is so low that we cannot assign this sample with certainty). As we don’t have 2 “Positive” genes, the sample is finally assigned as “Undetermined”.

b.2.11) The **ID_Result** tab displays a simple list of all analysed IDs and their final assignation. This file can be found in the toy dataset in folder **DYE-BioRad/output**. For this analysis, **ID_Result** tab must look like this:

| ID | Interpretation |
|---|---|
| 1 | Positive |
| 2 | Positive |
| 3 | Repeat |
| 5 | Positive |
| 6 | Negative |
| 7 | Repeat |

### Manual

##### 0) Before running your analysis

Before users run their own analysis, we first recommend to try out the Covid-19 toy dataset and to follow the manual provided in the app. Both the toy dataset and the manual can be downloaded by clicking the “Download Toy Dataset” and “Download Manual” buttons, respectively.

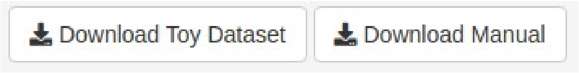


shinyCurves can be accessed in the following link: https://biosol.shinyapps.io/shinycurves/.

#### 1) Analysis choice

First, users have to choose the qPCR method used (**fluorescent probe (PROBE)** or **intecalating dye (DYE)** RT-qPCR assay) and system (**BioRad CFX** or **Applied Biosystems Quant Studio**). Data coming from any other qPCR system have to be adapted to the format of one of the sample files in the toy dataset.

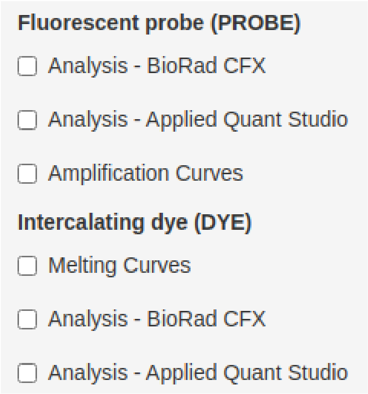


Upon clicking one of the “Analysis” options, a multi-option menu will be displayed in the grey box and a series of (empty) tabs will appear in the main panel on the right.

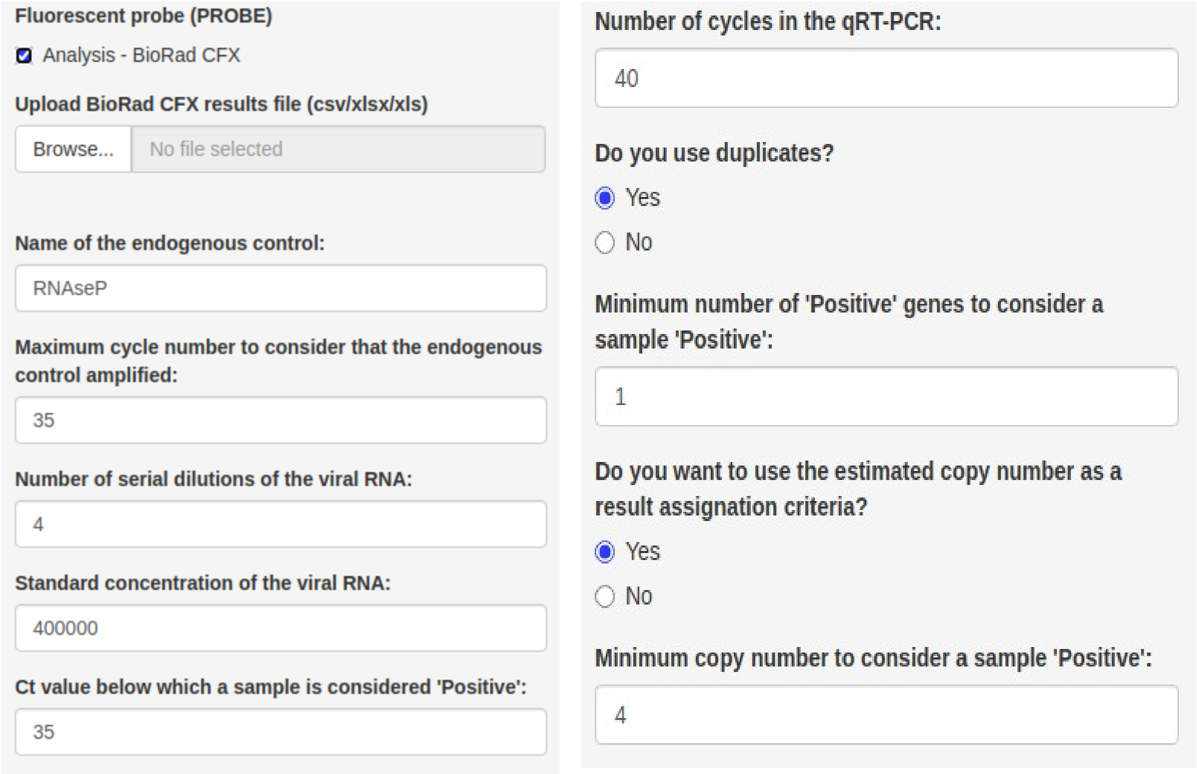

#### 2) File input and parameter choice

As a first step, users must load the data straight from the RT-qPCR system into the app and fine-tune several parameters. Analysis-specific data types and input file formats are described in the table below.

| Analysis | Required data | BioRad Format | AppliedFormat | Output files |
|---|---|---|---|---|
| ***Melting Curves*** | Temperature *vs* Cycle number | Melt Curve Results<br>- *csv* | Melt Curve Results<br>- *xlsx/xls*:<br>(**Melt Curve Raw Data** tab) | Melting Curve Plots (*pdf*)<br><br>Tm Table (*csv*)<br><br>ID_well (*csv*) |
| | ID<br>Target Gene<br>Well | Quantification Results<br>- *csv*<br>- *xlsx/xls*<br>(**Raw Data** tab) | Quantification Results<br>- *csv*<br>- *xlsx/xls*:<br>(**Results** tab) | |
| ***Calling*** | Well *vs* Ct | Quantification Results<br>- *csv*<br>- *xlsx/xls*<br>(**Results** tab) | Quantification Results<br>- csv<br>- xlsx/xls<br>(**Raw Data** and **Results** tabs) | Analysis table (*csv*)<br><br>ID_well (*csv*)<br><br>ID_result (*csv*) |
| | ID<br>Target Gene<br>Well | ID_well.csv | ID_well.csv | |
| ***Amplification Curves*** | RFU vs Cycle number | Quantification Amplification Results<br>- *csvs* | RFU adapted files | RFU adapted files<br><br>Amplification Curves Plots: all/ Undetermined samples (*png*) |
| | ID<br>Target Gene<br>Well | ID_well.csv | ID_well.csv | |
| | ID<br>Result | ID_result.csv | ID_result.csv | |

**a) Load Data**

shinyCurves accepts files directly exported from the qPCR system in different formats. Additionally, it is independent of the experimental setup, i.e. it will detect whether samples have been loaded into 96 or 364-well plates and the sample layout can be specified by the user, as long as the sample names follow a specific format (see “Input formatting” section)”.

- **Input formatting** – it is compulsory that sample names for dilutions, positive and negative controls coincide with the following format:

- Non-template-controls (NTC), usually $H_2O$, is used to detect potential contamination and is not expected to amplify: *NTC_gene* e.g. NTC_N1
- Negative controls (negC), commonly human genes, in which no amplification is expected either: *gene_negC* e.g. N1_negC
- Dilutions (if included) of the positive control (expected to amplify) and the target genes: *gene_10-dil* e.g. N1_10-5, RdRp_10-2
- *Note*: dilutions must compulsorily be 10-fold but non-consecutive points are allowed, i.e. 10-2, 10-3, 10-5.

Users must make modifications in the *Sample* column of their data files.

To upload the data files into the app, users must click the analysis-specific *Browse* button pictured below.

**a.1) BioRad CFX** - both for the PROBE and DYE analyses, users have different file upload choices:

**a.1.1)** (Required) Excel (*xlsx/xls*) file with a tab named **Data** (containing Ct Quantification Results). (Optional) Another tab in the same *xlsx/xls* file named **Run Information** (containing the run settings). Tab names must not be modified.

**a.1.2)** (Required) CSV file with the Ct Quantification Results. (Optional) Another CSV file with the Run Information.
*Note: Usually, these files have the common root name “Quantification Cq Results” or “Quantification Summary”, and “Run Information” after directly exporting the data from a BioRad platform.*

**a.1.3)** (Required-DYE ONLY – see Section 3 in this manual) *ID_well.csv* filewith *ID*, *Target* and *Well* columns. This file contains the IDs of samples that have shown acceptable melting curves in the **DYE-Melting Curves Analysis** and can be downloaded from the **ID_Well** tab resulting from the **DYE-Melting Curves Analysis.**

**a.2) Applied Biosystems Quant Studio**

**a.2.1)** (Required) Excel (*xlsx/xls*) file with 2 tabs named **Raw Data** (containing fluorescence data) and **Results** (containing Quantification Ct Results). Other tabs are allowed, but will not be taken into consideration.
*Note*: *commonly in Applied BQS machines, wells in which no amplification is detected are defined as Undetermined. In shinyCurves, these values will be converted to NAs.*

**a.2.2)** (Required-DYE ONLY – see Section 3 in this manual) Additionally for DYE Analysis, users must upload an *ID_well.csv* file with *ID*, *Target* and *Well* columns. This file must contain the IDs which have shown acceptable melting curves in the **DYE-Melting Curves Analysis**. This file can be downloaded from the **ID_Well** tab after performing **DYE-Melting Curves Analysis**.

> **IMPORTANT**: To differentiate wells in which no amplification is detected from wells in which no sample is charged, NA/NaN values in the Ct column for samples with **NO AMPLIFICATION** will be converted to 0 in the Analysis tab.

Upon successful loading of input data, the file-input bar should look like this:

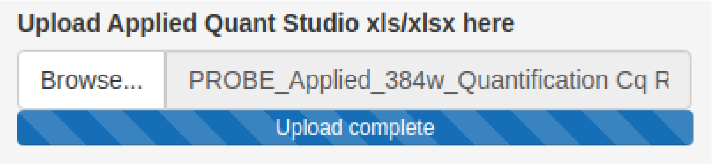


**a) Parameter Choice**

Users are allowed to modify several parameters:

❖ **Name of the endogenous control** (must written exactly as in the input files). *Default*: *RNAseP*

❖ **Maximum cycle number** to consider that the amplification for the **endogenous control** has been successful. *Default*: 35

❖ **Serial dilutions**: number of serial dilutions used in the standard curve. If 0 is chosen, the standard curve will neither be plotted nor used as a criterion for result assignation. *Default*: 4

❖ **Standard concentration**: number of viral nucleic acid molecules per µl contained in the stock control (copies/µl). This will be diluted as specified below to construct a standard curve for viral copy number estimation. *Default*: 400000

❖ **Ct value**, number of cycles equal to or below which a sample will be assigned as "Positive" (**MaxCt**). If the presence of amplification of a sample at any cycle will be considered as "Positive" then MaxCt = number of qPCR cycles. *Default*: 35

❖ **Maximum cycle number:** number of amplification cycles performed in the equipment. *Default*: 40

❖ **Duplicate** presence. *Default*: Yes

❖ When studying multiple genes, **number of necessary genes assigned as "Positive" to consider a sample "Positive" (#PosGenes)**. *Default*: 1

❖ **Use of estimated copy number as a result assignation criterion** in genes with Ct values falling between the **maximum cycle number to consider a sample "Positive"** and the **maximum cycle number**. *Default*: Yes

When "*No*" is chosen, samples will be assigned a result using only the Ct value cutoff criterion.

When "*Yes*" is chosen, users must fill another option:

- ❖ **Copy number** above which a gene will be assigned as "Positive" **(MinCopy#)**. *Default*: 4

#### 3) Melting Curves (DYE)

Before running DYE Analysis, users are allowed to plot **melting curves** for all samples in order to discard those which show unreliable melting temperature (Tm) peaks (indicating the presence of unspecific amplification products).
For this purpose, users must click the "*Melting Curves*" option in the DYE menu, which will display a new multi-option menu and 3 new tabs in the main panel.

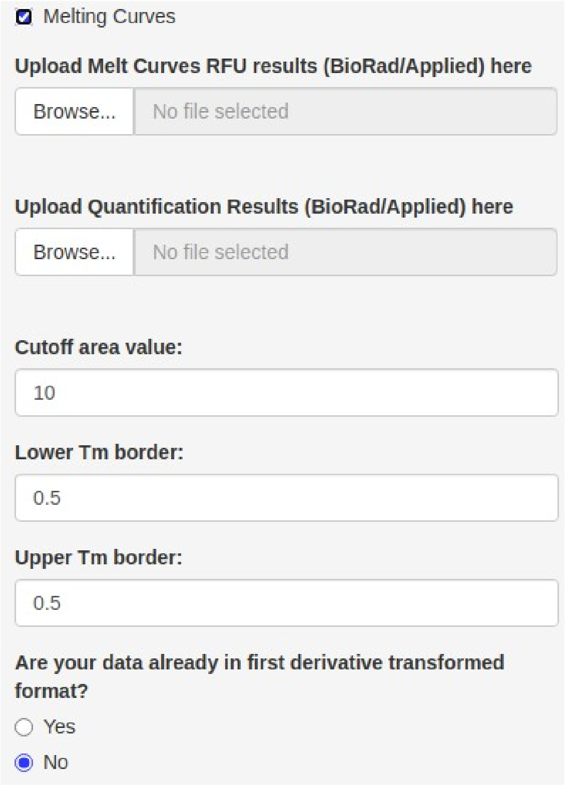


##### a) Load data

In this section, users must upload Melting Curve Result files and Quantification Results files.

###### a.1) BioRad CFX

**a.1.1) Melting Curve Results:** users must upload independent CSV files (one per gene).

**- Input formatting:** all filenames must have a common root name and end with "*_gene.csv*" (e.g. DYE_BioRad_96w_Melt Curve RFU Results_RdRp.csv). File content must not be modified.
*Note*: *Usually, these files have the common root name "Melt Curve RFU Results" after direct export from BioRad CFX platform.*

**- Upload:** to upload the data files into the app, users must click the analysis-specific "*Browse*" button shown below.

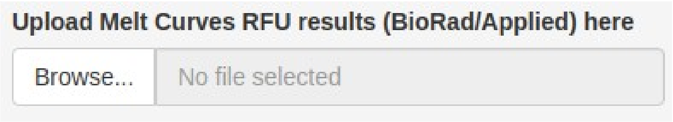

**a.1.2) Quantification Results**: users must upload the "*Quantification Results*" file in xlsx/xls format. Data will be read from the tab called **Data**.
The purpose of this file is to connect the independent wells from the "Melt Curve RFU Result" csv files (previously uploaded) with their respective Ct values in the "Quantification Results" file. Instead of the "Quantification Results" file, users are allowed as well to upload a "hand-made" *ID_Well.csv*, with columns *ID*, *Target* and *Well*.
*Note: After direct export from a BioRad CFX system, the tab in the "Quantification Results" containing Ct values is usually called "0". Users have to change this tab name to "Data".*

Upload this file using the button shown below:

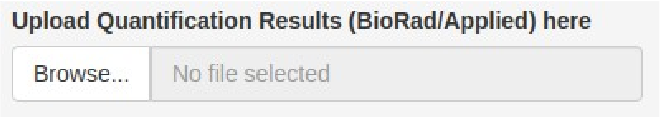


**a.2) Applied Biosystems Quant Studio**

**a.2.1) Melting Curve Results**: users must upload the output in xlsx/xls format. RFU data will be read from Melt Curve **Raw Data** tab. Tab names must not be modified.

**a.2.2) Quantification Results**: users must upload the same Applied output file in xlsx/xls format, but this time, data will be read from the **Results** tab. Tab names must not be modified.
The purpose of this file is to connect the independent wells from the "Melt Curve RFU Result" csv files (previously uploaded) with their respective Ct values in the "Quantification Results" file. Instead of the "Quantification Results" file, users are also allowed to upload a "hand-made" *ID_Well.csv*, with columns *ID*, *Target* and *Well*.

**b) Parameter choice**

Melting curves are plotted using the *meltcurve* function from the 'qpcR' *R* package (Spiess, 2018) and users are allowed to fine-tune several options as follows:
- *Cutoff area*: a peak area value to identify only those peaks with a higher area.
*Default*: 10
- *Lower and upper Tm borders*: for peak area calculation.
*Default*: 0.5 and 0.5
- *First derivative transformed format*: yes or no.
*Default*: Yes

**c) Melting Curves**

After successfully loading the input data and making the parameter choices, the following tabs will appear in the main panel:

- **ID_Well**: displays the ID_Well table (with columns *Well*, *Target* and *ID*) which is taken from the "Quantification Results" file.

- **Melting Curves Plots**: displays as many subtabs as genes analyzed. For each gene, a melting curve is plotted per well analyzed.
If users select "*No*" in the *First derivative transformed format* option, raw fluorescence data are plotted in black and the first derivative curve in red. The identified melting peaks are marked with a dotted vertical line (Spiess, 2018).

If users select "*Yes*" in the *First derivative transformed format* option, only the first derivative curve will be plotted in black. The melting peaks identified are marked with a dotted vertical line.
Melting curves can be downloaded in pdf format by clicking the "*Download PDF*" button.

- **Tm Table**: displays the Tm Table (*ID*, *Target*, *Well*, *Tm* and *Area* columns) for those wells showing unique peak area values above the cutoff.
The Tm Table can be downloaded in CSV format by clicking the "*Download CSV*" button.

- **New ID_Well**: similar to Tm Table but displaying only columns *ID*, *Target* and *Well*. This new ID_Well, which contains only those wells with acceptable and unique melting peaks, will be needed in the **DYE-Analysis** tab. It can be downloaded by clicking the "*Download CSV*" button.

#### 4) Calling analysis

##### a) PROBE/DYE Analysis

After successfully loading input data and adjusting parameters as specified in section 2, all tabs in the main panel should appear filled.

- **Raw Data**: displays the columns *Well*, *Fluor*, *Target*, *ID* and *Cq* read directly from input data.

- **Run Information**: displays run settings. If run information is not supplied, "*No run information to show*" will be printed.

- **Ct Plate**: displays a graphical representation of the plate with the Ct values read in each well. *Note: Ct = 0 represent empty wells.*

- **Sample Plate**: same as the Ct Plate, but displaying the name of the sample loaded in each well.

- (Optional) **Standard Curve**: if users have not included dilutions of all genes in the plate and do not want the standard curve to be plotted, they should choose 0 in the **Serial dilutions** option. In that case, the **Standard Curve** tab will only display the message "*No standard curve to show*".
On the contrary, if dilutions of all genes have been provided, this tab will display a table and a graphical tab as follows:

❖ **Controls/Dilutions Summary Table:**
<u>Fixed columns</u>
- *Dilutions*: as specified by the user.
- *Copies*: estimated copy number for each dilution using the formula Concentration/Dilution.
- *logCopies*: $\log_{10}$ of the estimated copy number (*Copies* column).

<u>Variable columns</u>
- (Duplicates) *Gene(Dup1)* and *Gene(Dup2)*: if users have included duplicates, each column will display the Ct value for each duplicate and dilution of the control target gene. Both columns will be displayed for all genes.
- (Duplicates) *Gene(Avg)*: average Ct value for the duplicates in each sample.
- (No Duplicates) *GeneCt*: Ct value for each dilution. This column will be displayed for all genes.

❖ **Standard Curve Plots:**
Independent standard curve plots for each of the target genes. The X-axis takes values from the *Gene(Avg)* column (when duplicates are present) and from the *GeneCt* column when they are not. The Y-axis always takes values from the *logCopies* column. The linear regression equation and the adjusted $R^2$ are shown.
*Note: dilutions without a Ct value are not included in the graph.*

- **Analysis**: displays a long table with the central Analysis of the app, in which samples are assigned a result ("Positive", "Negative", "Undetermined"). This table varies depending on the user's parameter choices. It can be downloaded by clicking the "*Download CSV*" button.

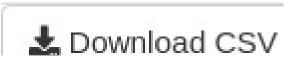

Samples are assigned a result by following the criteria represented in Supplementary Figure 1 and described below:

- ***Gene(MeanCt)/Gene(Ct)***: for all genes. If users have **included duplicates** and these do not differ significantly from each other (i.e. the absolute difference between Ct duplicates is smaller than 1.5), the mean Ct will be calculated and showed on column *Gene(MeanCt)*. If duplicates differ from each other (>1.5 Ct), shinyCurves will assign "Undetermined" to this sample, as duplicates will not be considered reliable. If users have **not included duplicates**, the abovementioned column will be replaced by *Gene(Ct)* and the Ct value for a given sample will be taken directly from the data file.
  *Note: wells with no amplification will show a 0, while wells with no sample will show NA.*

- ***CtCheck***: for each target gene. In this column, samples will be assigned a result based on their Ct value for that specific gene.

- If users have chosen **not to include the use of estimated copy number as a result assignation criterion:**

a) *Is the Ct value lower or equal to MaxCt and different from 0?*
**Yes**: the gene will be assigned as "Positive".
**No**: the gene will be assigned as "Negative".

- If users have chosen **to include the use of estimated copy number as a result assignation criterion:**

a) *Is the Ct value higher or equal to MaxCt and lower or equal to the maximum cycle number?*
**Yes**: gene will be assigned as "Check copy number".
**No**: see b)

*b) Is the Ct value lower or equal to MaxCt value and different from 0?*
**Yes**: gene will be assigned as "Positive".
**No:** gene is equal to 0 (no amplification) so it will be assigned as "Negative".

- ***Assignment/FinalCtCheck***: in this column, the independent *CtCheck* columns for all target genes are compared and each sample is assigned a final result (based exclusively on the Ct values).

  - If users have chosen **not to include the use of estimated copy number as a result assignation criterion**, this column will be called *Assignment*.

  a) *Is the number of "*Positive*" genes higher than #PosGenes?*
  **Yes**: sample will be assigned as "Positive".
  **No**: see b)

  b) *Is the number of "*Undetermined*" genes higher or equal to #PosGenes and higher or equal to the number of "*Positive*" and to the number of "*Negative*" genes?*
  **Yes***:* sample will be assigned as "Undetermined" (too many unreliable duplicates).
  **No***:* see c)

  c) *Is the number of "*Negative*" genes higher than the number of "*Positive*" genes and higher than the number of "Check copy number" genes?*

**Yes:** sample will be assigned as “Negative”.
**No:** sample will be assigned as “Undetermined”.

- If users have chosen **to include the use of estimated copy number as a result assignation criterion** this column will be call *FinalCtCheck*.

a) *Is the number of “*Check copy number*” genes higher or equal to #PosGenes and is it higher or equal to the number of “*Positive*” genes?*
***Yes***: sample will be assigned as “Check copy number”.
**No**: b)

b) *Is the number of “*Check copy number*” genes lower than #PosGenes and higher or equal to the number of “*Positive*” and “*Negative*” genes (independantly), and different from 0?*
***Yes****:* sample will be assigned as “Check copy number”.
***No****:* see c)

c) *Is the number of “*Positive*” genes higher than #PosGenes?*
**Yes:** sample will be assigned as “Positive”.
**No:** see d)

d) *Is the number of “*Undetermined*” genes higher than #PosGenes and higher or equal to the number of “*Positive*” and “*Negative*” genes?*
**Yes**: sample will be assigned as “Undetermined”.
**No**: see e)

e) *Is the number of “*Negative*” genes higher than the number of “*Positive*” genes and higher than the number of “*Check copy number*” genes?*
**Yes:** sample will be assigned as “Negative”.
**No:** sample will be assigned as “Undetermined”.

At this point, the analysis will be finished for users choosing not to use the estimated copy number as a result assignation criterion. The following columns will only appear when that option is chosen.

- ***LogCopies***: estimated copy number using Ct values and the gene-specific standard curve (formula: Ct * Slope + Coefficient).

- ***Copies***: 10^*LogCopies* column.

- ***CopyCheck***: for each target gene. In this column, samples will be assigned a result based on their estimated gene copy number.

  *a) Is the copy number higher or equal to MinCopy#?*
  **Yes**: gene will be assigned as “Positive”.
  **No**: gene will be assigned as “Undetermined”.

- ***FinalCopyCheck***: in this column, the independent *CopyCheck* columns for all target genes are compared and each sample is assigned a final result (based on the estimated gene copy number).

a) *Is the number of “*Undetermined*” genes higher or equal to #PosGenes and to the number of “*Positive*” genes?*
**Yes:** sample will be assigned as “Undetermined”.
**No**: see b)

b) *Is the number of “*Positive*” genes higher than #PosGenes?*
**Yes**: sample will be assigned as “Positive”.
**No**: sample will be assigned as “Undetermined”.

- ***Assignment***: this column takes the final assignation results for each sample respectively from the *FinalCtCheck* and the *FinalCopyCheck* columns (depending on which criteria have been used to assign a result for each sample).

  - **ID_Well**: displays a table with columns *Well*, *Target* and *ID* for all wells, which can be downloaded by clicking the “*Download CSV*” button.

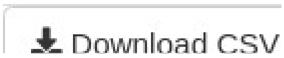


  - **ID_Result**: displays a table with columns *ID* and *Interpretation* for all samples. The *Interpretation* column is filled with the *Assignment* column of the Analysis tab. This table can be downloaded by clicking the “*Download CSV*” button.

  *Note: these files will be needed to perform the* ***Amplification Curves*** *analysis in the PROBE analysis.*

#### 5) Amplification Curves (PROBE)

After completing the PROBE Analysis, users may plot the **amplification curves** for all samples. For this purpose, they should click the "*Amplification Curves*" option in the PROBE menu, which will display a new multi-option menu and 4 new tabs in the main panel.

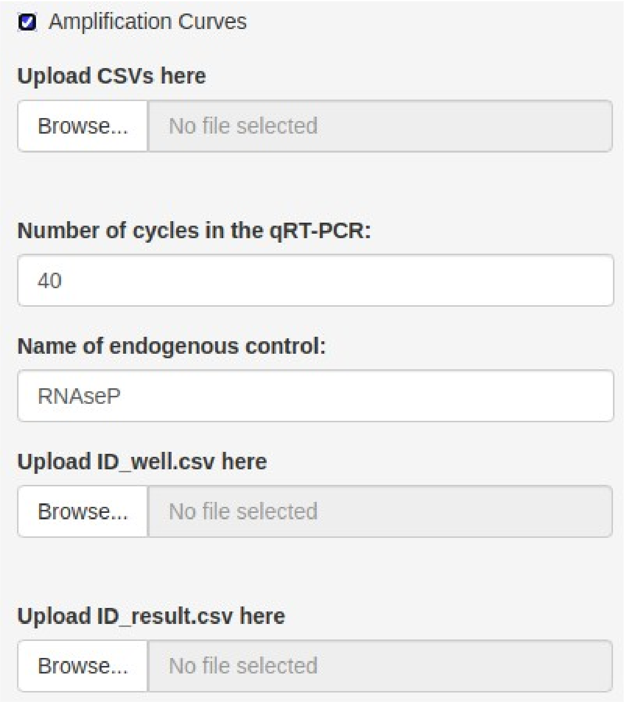


##### a) Load Data

In this section, users have to upload independent files with the amplification results for each gene.

**a.1) Quantification Amplification RFU Results**: users must upload independent CSV files (one per gene).

**- Input formatting:** all filenames must have a common root name and end with "*_gene.csv*" (e.g. PROBE_BioRad_364w_Quantification Amplification Results_N1.csv) or simply be named "*gene.csv*" (e.g. N1.csv, RdRp.csv). The file content must not be modified.
*Note: Usually, these files have the common root name "Quantification Amplification Results" after direct export from a BioRad CFX platform.*
*Note: When analyzing Applied BQS data, these files can be obtained in the* ***Conversion*** *tab in the Analysis-Applied Biosystems Quant Studio option.*

**- Upload:** to upload the data files into shinyCurves, users must click the analysis-specific "*Browse*" button shown below.

Additionally, users must upload the *ID_well.csv* and *ID_result.csv* files obtained from the main analysis (Analysis - BioRad CFX/Applied Biosystems Quant Studio). These must be uploaded into the cells shown below, respectively.

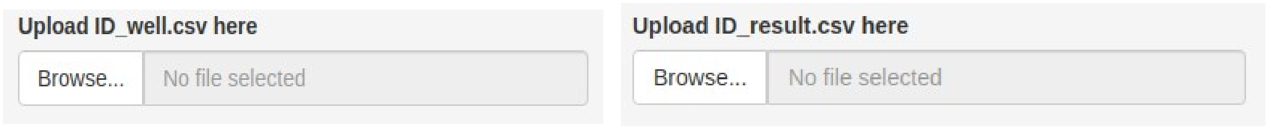


**- Others:** users must also specify:
- Name of the endogenous control. *Default*: *RNAseP*
- Number of cycles performed in the RT-qPCR. *Default*: 40

**b) Amplification Curves**

After successfully loading the input data, the following tabs will appear in the main panel:

- **ID_Well**: displays the ID_Well table directly from the uploaded file.

- **ID_Result**: displays the ID_Result table directly from the uploaded file.

- **General Plots**: displays the amplification curves of all the analyzed samples for each gene. The X-axis represents the cycle numbers (Ct) while the Y-axis corresponds to the RFU data. These plots are generated by the R package ‘plotly’ (Sievert, 2020) and are interactive, which means that, when the mouse is passed over a curve, a label indicating the Sample ID, Well, RFU data and Interpretation of that sample will be displayed.
Samples assigned as “Positive” are represented in green, those assigned “Negative” in red and “Undetermined” samples in blue.
Amplification curves can be downloaded in png format by clicking the camera button on the upper right side of the plot.

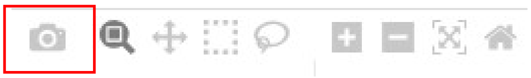

- **Undetermined Samples**: displays as many subtabs as genes analyzed. Each tab shows the individual amplification curves for each sample that has been assigned as “Undetermined”, together with all the “Positive” and “Negative” samples. If no “Undetermined” samples are present in an experiment, these tabs will display the message “*No Undetermined samples to plot*”.
Amplification curves can be downloaded in png format by clicking the thecamera button on the upper right side of the plot.
