## Supplementary figures and images for "shinyCurves, a shiny web application to analyse multisource qPCR amplification data: a COVID 19 case study"

### Supplementary Figure 1

#### Supplementary Figure

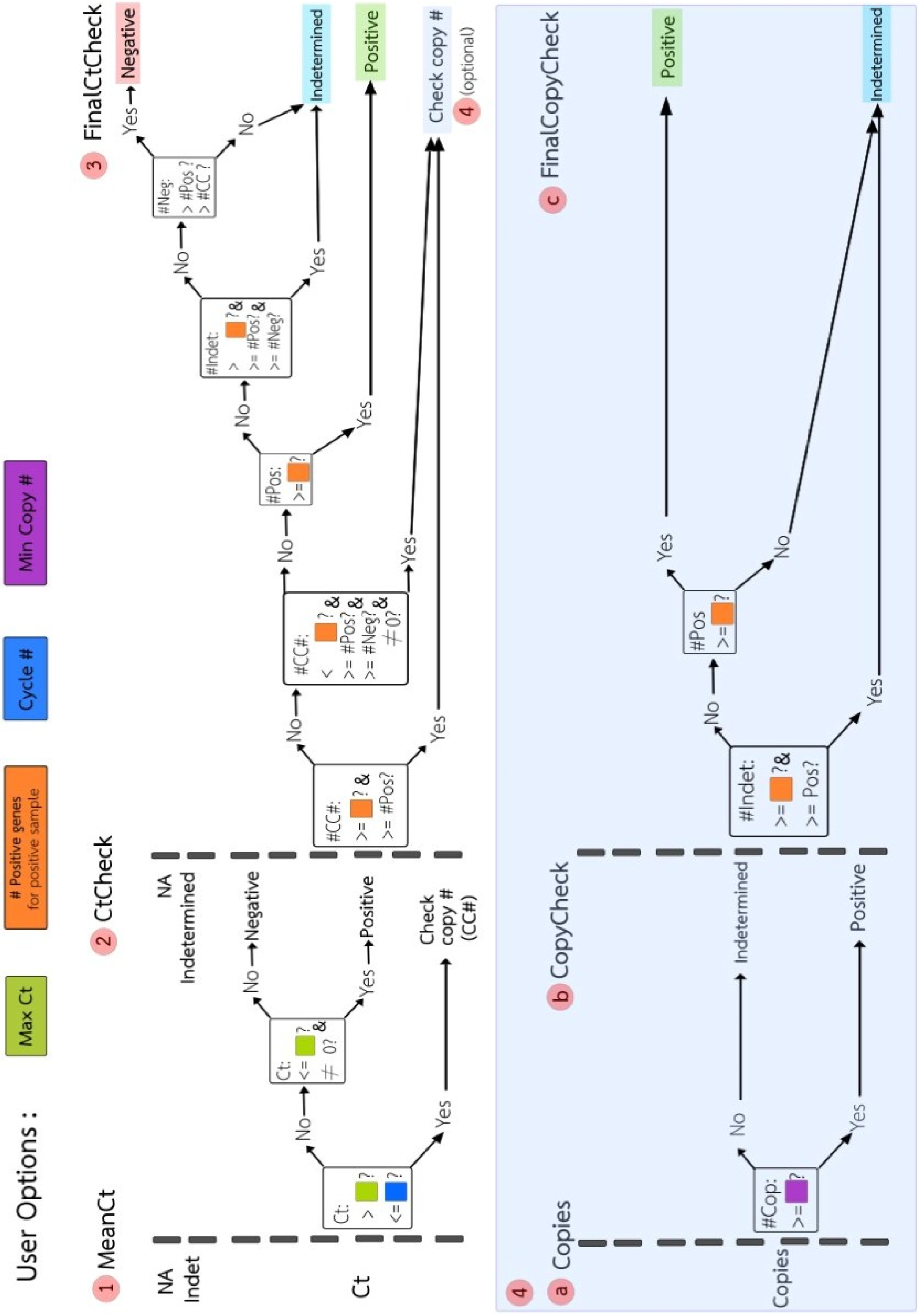
